# Qualia NAD+® Increases NAD+ Levels In Human Participants: A Double-Blind, Placebo-Controlled Study

**DOI:** 10.1101/2025.03.19.25324259

**Authors:** Sarah A. Blomquist, Gregory Kelly, Sara Adães, Christopher R. D’Adamo, Abhimanyu Ardagh, Shawn Ramer, William Scuba

## Abstract

**Background:** Nicotinamide adenine dinucleotide (NAD) is a crucial coenzyme involved in cellular energy homeostasis whose levels decline notably with aging, which has prompted interest in NAD+ boosting to help combat age-related diseases and dysfunction. Numerous clinical trials have demonstrated safety and efficacy for B3 vitamins, such as nicotinamide riboside (NR), to increase NAD+ levels and augment the NAD+ metabolome. Limited impact on clinically relevant outcomes or quality of life have been demonstrated, however.

**Objective:** This double-blind, placebo-controlled, remote pilot study examined the effects of supplementation with Qualia NAD+® on whole blood NAD+ levels and quality of life measures.

**Methods:** Study participants consumed Qualia NAD+ daily for 28 consecutive days. NAD+ levels were the primary outcome and measured using a self-administered, non-invasive blood spot assay test conducted at baseline and at study end. Quality of life questionnaires were the secondary outcomes and reported bi-weekly. NAD+ levels and questionnaire data were analyzed using a linear mixed-effects model to compare within-group and between-group differences over time as compared to baseline. Independent t-tests were utilized to compare safety and tolerability between study groups.

**Results:** Twenty-five healthy adults aged 40 - 65 (56% female) consumed Qualia NAD+ (n = 9) or placebo (n = 16) Overall, Qualia NAD+ significantly increased NAD+ levels an average of 74% compared to a 4% increase observed in the placebo group (p < 0.001). Within-groups comparisons for Qualia NAD+, and changes in NAD+ levels comparing Qualia NAD+ and placebo were also significant (p < 0.001). Improvements in the overall and somatic categories (p = 0.02 and p = 0.04) were observed for males only comparing baseline to the end of the study. No adverse events were reported.

**Conclusions:** Qualia NAD+, a novel nutraceutical formulated with multiple vitamins, nutrients, and botanical compounds, effectively increased whole blood NAD+ levels and may improve some symptoms of aging in males. Clinicaltrials.gov identifier: NCT06812416.

## Introduction

Since the discovery of nicotinamide adenine dinucleotide (NAD[H]) over a century ago^1^ it has received the reputation as a universal energy- and signal-carrying molecule, as well as a super-centenarian molecule. Besides its essential role as a coenzyme in cellular oxidation-reduction (redox) reactions that are fundamental to almost all metabolic pathways, NAD(H) levels decline during chronological aging in both animals^2,3^ and humans^4-10^, and in age-related conditions like diabetes, cardiovascular and neurodegenerative diseases^11-17^. The finite pool of NAD+ is also under competition for use by NAD+-dependent enzymes including poly-ADP-ribose polymerases (PARPs), sirtuins (SIRT) and cluster of differentiation (CD38) in DNA repair, cellular signaling, immunity and inflammation, respectively ^18-21^. These roles of NAD+ are involved in the modulation of healthspan and lifespan^22-26^. With aging, these NAD+-dependent enzymes face increased stress, leading to a higher demand for resources like NAD+^3,4,27-29^. For these reasons, there has been tremendous interest in boosting NAD+ levels. Emerging preclinical and clinical evidence suggests this may mitigate age-related disease and dysfunction^30-34^.

NAD is an organic cofactor synthesized via the *De Novo* Pathway starting from the essential amino acid L-tryptophan, via the Preiss-Handler Pathway using the B3 vitamin nicotinic acid, via the Salvage Pathway using the B3 vitamin nicotinamide riboside (NR), and via the Reduced Salvage Pathway starting with the reduced forms of NR and nicotinamide mononucleotide (NMN) (**Figure 1**). Over a dozen clinical trials have examined approaches to increase NAD+ and associated health outcomes, primarily by administering different forms of B3 vitamins that enter the Salvage pathways. For example, in a randomized controlled trial (RCT) with 133 healthy overweight adults aged 40 - 60, consumption of 100, 300 and 1000 mg Niagen® NR and dose-dependently and significantly increased whole blood NAD+ by an average of 22%, 51% and 142%, respectively, within 2 weeks; increases were sustained up to 8 weeks^35^. Studies have collectively demonstrated the safety of the NAD+ boosters NR and NMN when administered at various doses for periods of up to 20 weeks ^36^, while nicotinic acid has been administered for up to 6 years ^37^.

**Figure 1:**
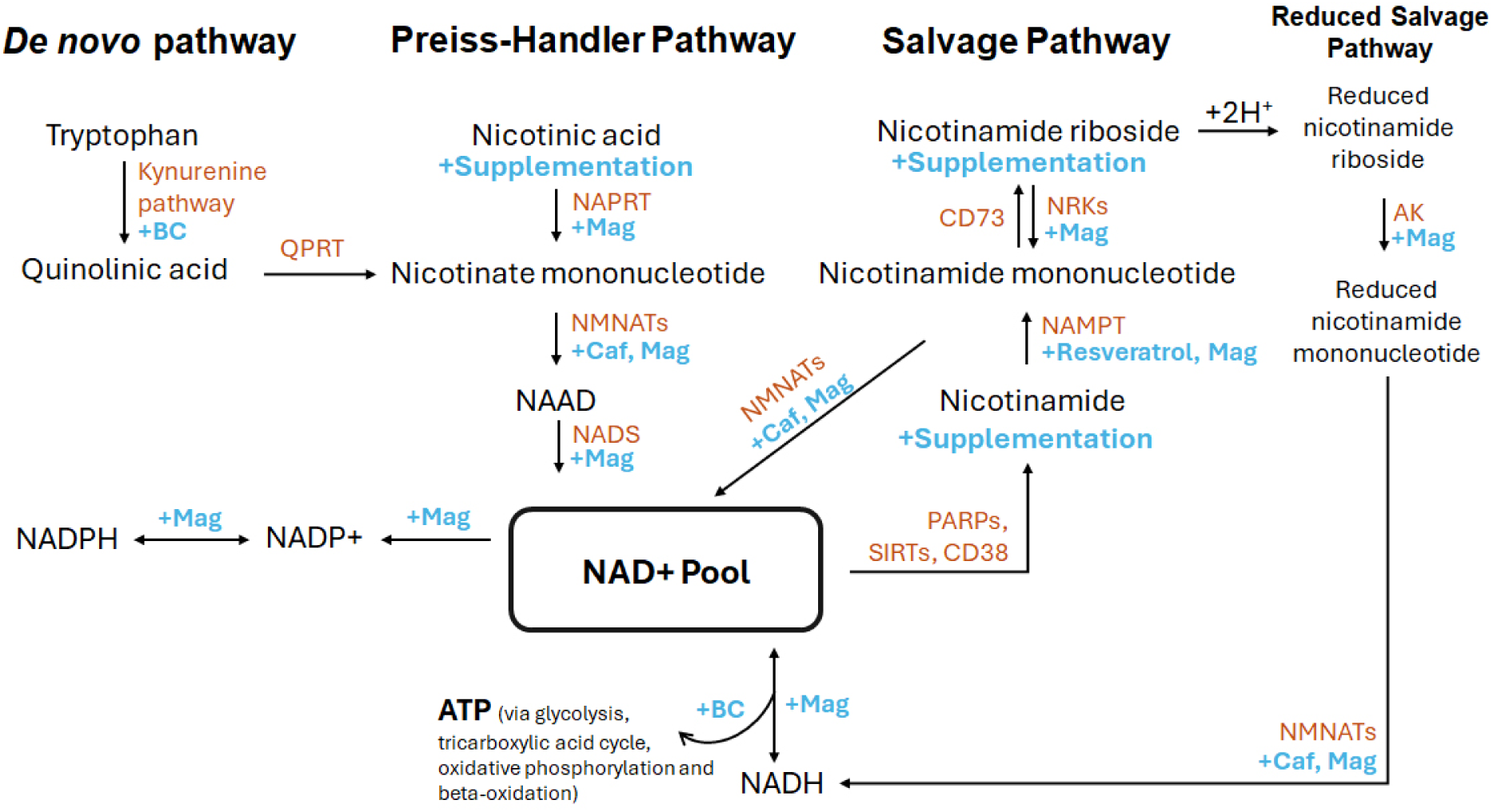
Hypothesized Targets of Qualia NAD+ Supplementation Within NAD+ Metabolic Pathways. Enzymes are colored orange and pathway support by Qualia NAD+ is colored blue and designated by + and the compound, nutrient, or “Supplementation”, meaning the B3 vitamin itself was provided. AK: adenosine kinase; BC: Vitamin B complex; Caf: Caffeine, CD38: Cluster of differentiation 38; Mag: Magnesium; NAAD: nicotinic acid adenine dinucleotide; NADS: NAD+ synthase; NAMPT: nicotinamide phosphoribosyltransferase; NAPRT: nicotinic acid phosphoribosyltransferase; NMNATs: nicotinate N-methyltransferases; NRKs: nicotinamide riboside kinases; PARPs: poly (ADP-ribose) polymerases; QPRT: quinolinate phosphoribosyl transferase; SIRTs: sirtuins

In its redox role, NAD(H)’s oxidized form, NAD+, accepts high-energy electrons to form NADH, which then donates the electrons to the electron transport chain (ETC) for adenosine triphosphate (ATP) synthesis. Shifting from NAD+ to NADH is essential for catabolic processes such as glycolysis, fatty acid oxidation, and the citric acid cycle. NAD+ can also acquire a phosphate on the adenine ribose to form NADP+. The interconversion between oxidized NADP+ and reduced NADPH drives anabolic processes like fatty acid synthesis, cholesterol, and nucleic acid production. NAD+/NADH and NADP+/NADPH ratios are central in regulating metabolism and may be more important than the amount of NAD+ in isolation, underscoring the importance of also considering the impact of the broader NAD+ metabolome in age-related disease and dysfunction^6,38,39^.

Studies combining B3 vitamins with additional nutrients or ingredients have also revealed promising modulation to the broader NAD+ metabolome and clinical biomarkers. A RCT examined the consumption of nicotinamide and D-ribose (RiaGev®) in 18 healthy adults aged 35 - 65 and observed that 1520 mg taken twice daily for 7 days significantly increased NAD+ by ∼10% and NADP+ by ∼27% at day 5^40^. Improvements in glutathione, blood glucose, waking salivary cortisol, and quality of life measures were also noted. Another RCT administered Nuchido TIME+® (500 mg nicotinamide plus other nutrients and ingredients) in 26 healthy adults aged 21 - 72 and observed increased NAD+ concentrations in whole blood by an average of 26.5%. They also noted increases in SIRT1 and nicotinamide phosphoribosyltransferase (NAMPT) in peripheral blood mononuclear cells (PBMCs), reduced pro-inflammatory cytokines and glycated serum protein, and a shift in the glycosylation profile of immunoglobulin G toward a younger biological age^41^. Overall, B3 supplementation effectively boosts NAD+ levels and augments the NAD+ metabolome, but its impact on clinical outcomes and quality of life is still limited (studies reviewed in more detail by Freeberg et al^36^).

The product under study, Qualia NAD+, was developed using a complex systems science approach with the intent to holistically support NAD+ metabolism by (i) delivering multiple B3 vitamers (NR, nicotinic acid, nicotinamide) that rely on different pathways and/or enzymes before converging on NAD+, and (ii) enhancing the activity of these NAD+-synthesizing enzymes using a combination of a magnesium, resveratrol, other B vitamins, and a coffee fruit extract standardized for caffeine (**Figure 1**). Therefore, the objective of this hypothesis-generating pilot study was to examine the impact of Qualia NAD+ on changes in (i) NAD+ levels as measured by a self-administered, non-invasive blood-spot assay card, and (ii) quality of life measures. Although NAD+ and its metabolites exhibit stability challenges when measured in various biological tissues^6,42^, NAD+ may be stabilized by a treated assay card^42-45^. Additionally, while we recognize that the NAD+/NADH ratio is a more comprehensive indicator of cellular redox status, this study focused on measuring blood NAD+ as an initial step in assessing the biological impact of the supplement. To test our objective, we gave healthy adults aged 40 - 65 either Qualia NAD+ or a placebo for 28 consecutive days, then compared baseline measurements of whole blood NAD+ levels and quality of life survey scores to their respective end values.

## Methods

### Study product

Qualia NAD+ (Qualia Life Sciences, Carlsbad, CA, USA) is a novel dietary supplement formulated to support the maintenance and enhancement of NAD+ levels. The serving size of two capsules contains the following: 0.32 mg vitamin B1, 0.4 mg vitamin B2, 254 mg of vitamin B3, (as 234 mg niacinamide [also nicotinamide] and 20 mg niacin [nicotinic acid]), 0.49 mg vitamin B6, 166 mcg dietary folate equivalents (DFE) folate, 1.5 mcg vitamin B12, 37.5 mcg biotin, 1.375 mg vitamin B5, 50 mg magnesium (as Aquamin® magnesium hydroxide from seawater [Marigot Ltd., County Cork, Ireland]), 300 mg Niagen® (nicotinamide riboside chloride from Chromadex Corp., CA, USA), 150 mg trace mineral blend (Aquamin®), 50 mg trans-resveratrol, and 28 mg caffeine from organic Coffeeberry® (Futureceuticals, USA) whole coffee fruit extract. All of these ingredients meet the criteria for dietary supplements established by the FDA, and all dosages are safe.

### Study design and intervention

The current study is a double-blind, placebo-controlled, parallel-arm trial. The trial was registered at ClinicalTrials.gov (identifier: NCT06812416). All participants were assigned in a 1:1 allocation ratio, stratified based on age and gender, to receive a 28-day supply of either Qualia NAD+ or placebo (Nu-Flow®, rice powder in a cellulose/vegetable capsule) to be taken daily in the morning, with or without food. The primary outcome was to investigate the efficacy of Qualia NAD+ in increasing NAD+ levels in blood samples at the end of study compared to baseline and placebo, and the secondary outcomes were findings from health-related quality of life questionnaires.

### Blinding

To ensure that all subjects and researchers were unaware of the treatment assignments, a Qualia Life Sciences staff member unassociated with the research team labeled supplement bottles A or B. The research team was blinded to the content of the bottles and the information was kept confidential until study conclusion. In the event of an emergency or if the participant decided to not participate, their condition was revealed.

### Participants

Participants were recruited in 2 separate cohorts via an online recruitment platform to conduct this fully remote clinical trial. Interested participants completed a prescreen survey to determine eligibility. Adults who met the inclusion criteria were contacted to discuss the intervention and obtain informed consent. Subjects were included if they were healthy male and female participants between the ages of 40 - 65 and were willing to: provide a valid cell phone number and receive text communications, complete questionnaires associated with the study, abstain from supplements or products containing vitamin B3 four weeks prior to baseline and through the intervention, continue taking any supplements they regularly use (with the exception of those containing vitamin B3) during the study period, and self-administer the NAD+ fingerstick test at baseline and end of study. The exclusion criteria included: women who were pregnant, breastfeeding, or planning to become pregnant during the trial, known food intolerance/allergy to any ingredients in the product, record of significant cardiovascular events in the last 6 months, currently taking MAO inhibitors, SSRIs, or any other psychiatric or neurological medicines, currently on immunosuppressive therapy, those with psychiatric conditions, neurologic disorders, endocrine disorders, or cancer, or those who were incompatible with the test protocol or unable to provide consent.

### Ethics

The study protocol was approved by the Advarra Institutional Review Board (protocol number: Pro00073942) and adheres to international ethical standards for human research, including the principles outlined in the 2024 Declaration of Helsinki. Participants provided written informed consent before data collection. All authors were offered full access to the study data and reviewed and approved the final manuscript.

### NAD+ levels testing

Revvity (previously part of PerkinElmer) developed a clinical assay to measure whole blood NAD+ levels from dried blood spot cards (DBS). Samples are self-collecting by fingerstick using a lancet on chemically coated DBS cards. DBS assessment of NAD+ has been validated previously ^43,46^. Participants were given standardized instructions, and blood spot samples were collected individually by each participant, then dried and stored at room temperature (15 – 30°C). Participants were instructed to complete their blood test and samples were shipped within seven days of collection. Upon arrival at the laboratory, samples could receive analysis on the same day, otherwise samples were stored at ≤ − 20 °C at the laboratory until processing. The NAD+ assay measurements use an assay that uses an externally calibrated isotope dilution protocol, a heavy labeled internal standard, and liquid chromatography/tandem mass spectrometry (LC-MS/MS). The analyses were conducted at a CLIA-/CAP-accredited clinical genomics and biochemical laboratory.

### Aging Males Symptoms Scale

The Aging Males’ Symptoms (AMS) Scale is a validated questionnaire designed to assess symptoms of health-related quality of life and severity of symptoms in aging males over time^47^. The questionnaire consists of 17 items grouped into 3 categories: psychological, somato-vegetative, and sexual. Each item is scored on a severity scale ranging from 1 to 5, where higher scores indicate more severe symptoms. While it can be used in males of all ages, it was designed with men over 40 in mind and is used internationally for clinical research. Participants were asked to complete this survey bi-weekly (baseline, days 14 and 28).

### Aging Females Symptoms Scale

The AMS scale was modified for this study to create a version that could be used with females, however most of the questions are gender-neutral and can be used for all participants. Questions 14-16 in the AMS were male-specific, therefore question 14 has been modified to produce a female version and questions 15 and 16 were eliminated. The same scoring and severity scale were applied as in the AMS. Participants were asked to complete this survey bi-weekly (baseline, days 14 and 28).

### RAND-36

The 36-Item Short Form Survey (SF-36) was developed by RAND as part of the Medical Outcomes Study^48,49^. Questions are grouped into eight subscales intended to assess physical functioning, bodily pain, role limitations due to physical health problems, role limitations due to personal or emotional problems, emotional well-being, social functioning, energy/fatigue, and general health perceptions. It also includes a single item that provides an indication of perceived change in health. Each item is scored on a 0 to 100 range so that the lowest and highest possible scores are 0 and 100, respectively. Scores represent the percentage of total possible scores achieved. A high score defines a more favorable health state. Participants were asked to complete this survey bi-weekly (baseline, days 14 and 28).

### Safety and tolerability

Participants were asked to complete 9 questions in a self-reported daily diary to assess adherence and side effects. They were also asked to record how many capsules they took, the time of day they were taken, and whether the capsules were taken with food. On a weekly basis, participants were also asked to complete a safety and tolerability survey. The possible responses for the safety and tolerability survey were: not at all, a little, a moderate amount, a lot, and a great deal, with scores corresponding from 0-4, respectively.

### Statistical analysis

Safety and tolerability data were analyzed in Python using an independent t-test to compare between groups and a dependent t-test to assess changes within groups from baseline to respective end points (days 14 and 28). NAD+ blood levels and survey data were analyzed in Python using a linear mixed-effects model (LMM) to compare within-group and between-group differences over time compared to baseline. The model included fixed effects for group (Qualia NAD+ vs. placebo), time (baseline, days 14, 28), and the interaction between group and time, as well as a random effect for participants to account for repeated measurements within individuals. The models were fitted using a maximum likelihood estimation method to optimize parameter estimates. Effect sizes (reported as Cohen’s *d*), confidence intervals (calculated from a paired *t* confidence interval), and point estimates (reported as raw β coefficient) are reported for between-groups comparisons. For the LMM, total standard deviation was used to capture both within and between-groups variation. A modified per-protocol analysis (mPP) was reported as the primary results, where participant data was excluded from the analysis for the following reasons (i) the participant indicated taking <80% of the product dosage over the trial period and (ii) the data were recorded outside the designated window of time for that assessment. A per-protocol analysis (PP) was also utilized to examine the survey results, where only participants who completed baseline and end of study bloodwork were included. An intention to treat (ITT) analysis was also performed, where all participants allocated to Qualia NAD+ or placebo were included regardless of compliance or attrition. The sample size was estimated for the study design, yielding 80% power to detect an effect (Cohen’s d = 0.80) with a two-tailed t-test at a 0.05 significance level.

## Results

### Study population

Participants were recruited in 2 separate cohorts between September - October 2023 and during January 2024. This study was conducted between October 2023 and March 2024. Ninety-two adults were recruited and sent at-home finger stick blood kits to measure initial blood NAD+ levels prior to beginning supplementation.

Forty-seven participants successfully completed the initial test to obtain their baseline NAD+ measurements and were instructed to take Qualia NAD+ or placebo. Twenty-five of the 47 participants (51.0%) successfully completed a second NAD+ test after taking Qualia NAD+ or a placebo for 28 days (**Figure 2**). The participants who had both initial and final NAD+ measurements were aged 40 to 64 years old with an average age of 49.5 years. At baseline, no significant differences comparing NAD+ levels in the Qualia NAD+ and placebo groups were observed (p = 0.73). Participants were a mix of females (n = 14) and males (n = 11). Data analysis using the mPP analysis for NAD+ levels is based on these 25 participants who had both initial and final NAD+ measurements. The most common reasons for failing to successfully complete NAD+ measurements were: (i) failure to send in the initial or final at-home finger stick blood kits to the lab for analysis, (ii) insufficient blood sample of the submitted test kit, and (iii) too much time having elapsed between when the sample was collected and returned to the lab.

**Figure 2:**
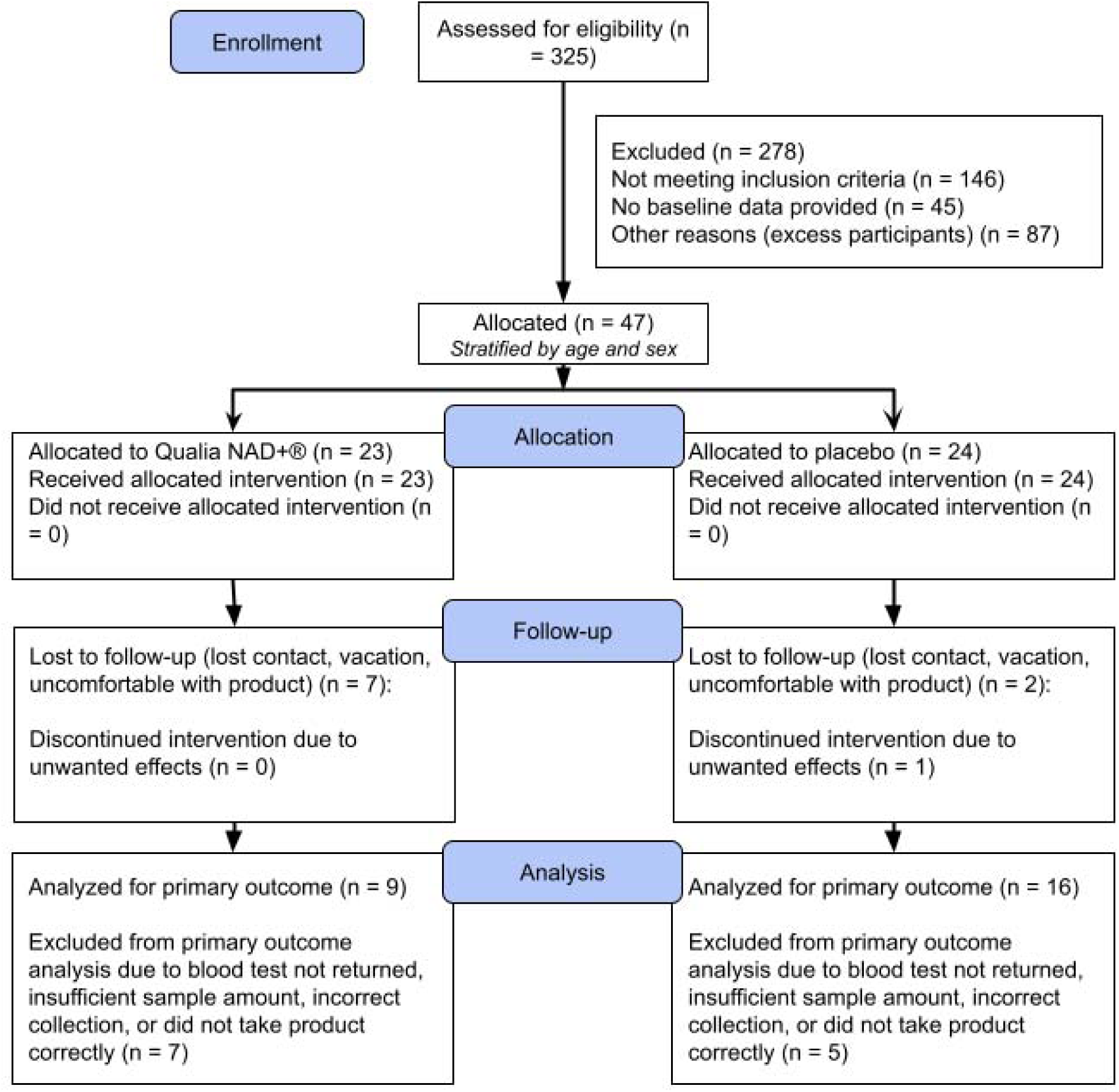
Participant flow chart.

Fifty-eight participants were included in the survey analysis for the mPP analysis. Reasons for excluding participants from the survey analysis are the following: (i) 15 participants did not meet the dosage threshold of 80% (described in methods), (ii) 18 participants only completed the baseline survey, and (iii) 6 participants did not complete any surveys. The twenty-five participants who completed both initial and final NAD+ measurements were also examined in a PP analysis for the survey data. For the ITT analysis, Fourty-four participants were included in the Qualia NAD+ group and 44 participants were included in the placebo group for both blood levels and survey data.

### Primary endpoint

In the mPP analysis, participants receiving Qualia NAD+ exhibited a mean increase in NAD+ levels of 14.56 µM (74.1%) while the placebo group experienced a mean increase of 0.71 µM (4%). Compared to baseline values, Qualia NAD+ significantly increased NAD+ (p < 0.001) while there was no change in the placebo arm (p=0.35) (**Table 1**). Both the final between-group differences (p < 0.001, Cohen’s *d* = 2.88, 95% CI [9.98, 18.88]), and the changes in NAD+ levels between groups were statistically significant at the study’s conclusion (p < 0.001, Cohen’s *d* = 3.31, 95% CI [10.2, 17.5]). Intraindividual variability in response to Qualia NAD+ supplementation was observed, with changes from baseline ranging from 29% to 134% (median 78%), as shown in **Figure 3**. The ITT analysis revealed similarly that when comparing Qualia NAD+ to placebo on day 28, there were significant increases in NAD+ levels (p < 0.001 Cohen’s *d* = 3.02, 95% CI [10.4, 16.5]).

**Figure 3:**
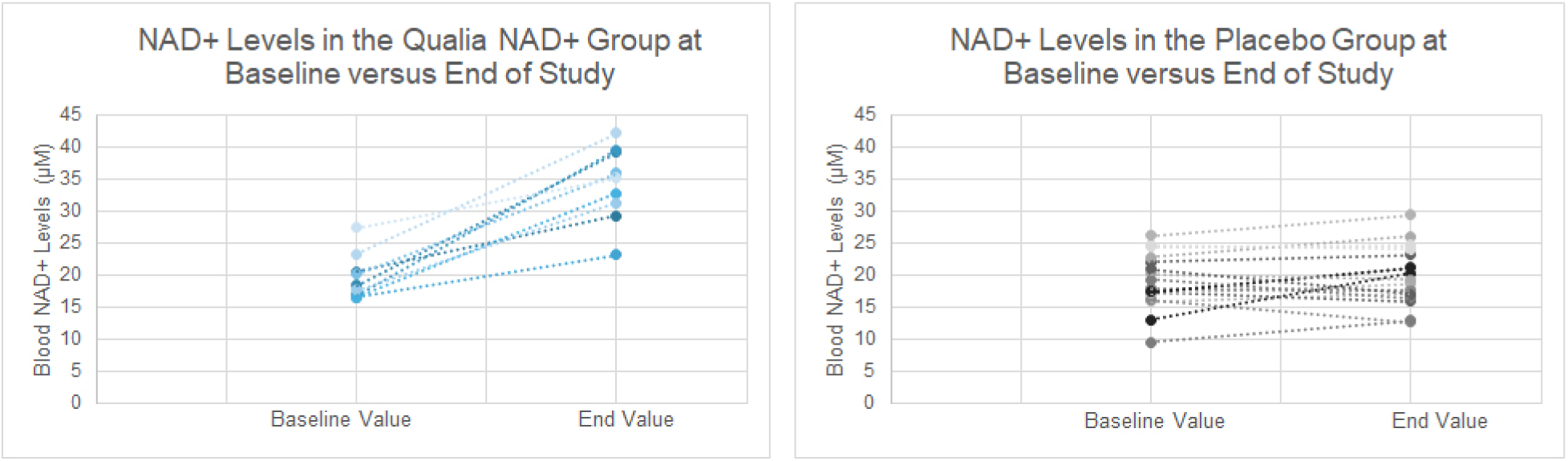
NAD+ Levels in the Qualia NAD+ and Placebo Groups. NAD values were measured using the self-administered NAD+ fingerstick at the baseline (beginning of the study) and end values were measured after 28 consecutive days of supplementation in the NAD+ group (n = 9) or the placebo group (n = 16). Solid dots connected with dotted lines represent baseline and end values for an individual participant.

**Table 1:** Descriptive Statistics for Group Comparisons. Means are shown with ± standard deviations. The change value is the end value (day 28) minus the baseline value. * p < 0.001 when compared to the placebo group.

| Group | n | Baseline Value<br>(μM) | End Value (μM) | Change (μM) | P-value<br>(within-group) |
| --- | --- | --- | --- | --- | --- |
| NAD<sup>+</sup> | 9 | 19.64 ± 3.63 | 34.20 ± 5.88* | 14.56 ± 5.79* | < 0.001 |
| Placebo | 16 | 19.06 ± 4.44 | 19.77 ± 4.70 | 0.71 ± 3.00 | 0.35 |

### Secondary endpoints

In the mPP analysis, significant improvements were observed in the between-groups differences for the overall (p = 0.02, β = -4.61, Cohen’s *d* = 0.93, 95% CI [0.586, 8.651]) and somatic (p = 0.04, β = -2.16, Cohen’s *d* = 0.76, 95% CI [0.073, 4.261]) categories for the Aging Male Symptom Scale (AMS) when comparing baseline to day 28 (p-values are not adjusted for multiple comparisons). No significant improvements were observed comparing baseline to days 14 or 28 for the Aging Female Symptom Scales (AFS) or the 36-Item Short Form Survey (RAND-36) (all p > 0.05, data not shown).

In the PP analysis for the AMS, there were significant improvements in the NAD+ group compared to placebo in overall (p = 0.04), sexual (p = 0.02), and borderline for somatic (p = 0.054) at day 14 compared to baseline. For the RAND-36, significant improvements in energy/fatigue (p = 0.003), emotional well-being (p = 0.002) and social functioning (p = 0.01) were observed on day 28 compared to baseline in the NAD+ group compared to placebo. In the ITT analysis, no significant differences between-groups were observed when comparing baseline to days 14 or 28 for the AMS, AFS, or RAND-36.

### Safety and tolerability

No significant differences were observed between NAD+ and the placebo groups in the safety and tolerability assessment (p > 0.05) when comparing baseline scores to days 14 and 28, indicating Qualia NAD+ was well tolerated by participants (**Figure 3**). In the placebo group, only one individual dropped out of the study due to reported side effects.

**Figure 4.**
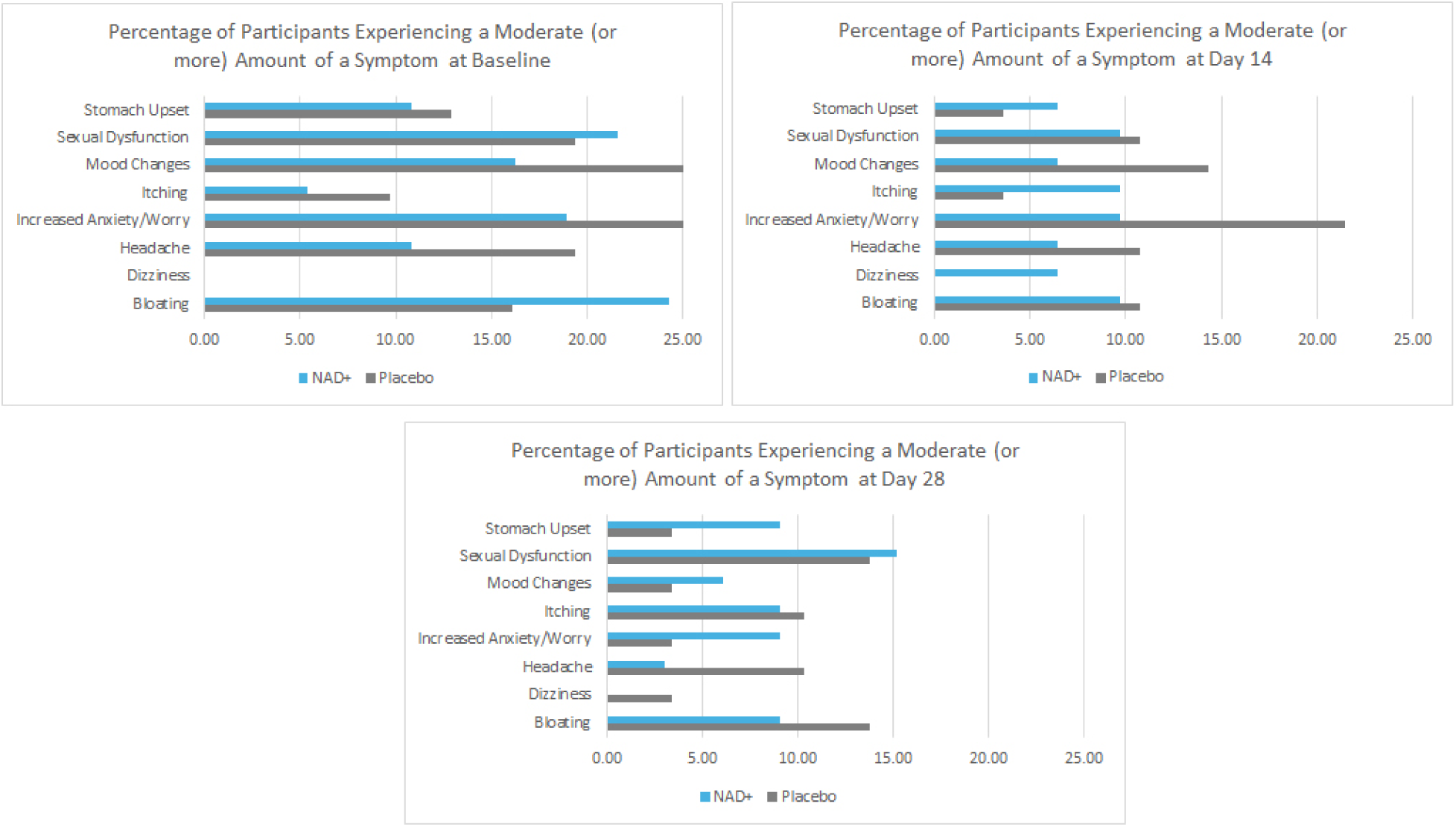
Percentage of Patients Experiencing Symptoms Comparing NAD+ and Placebo Groups. Data is from the mPP analysis. Moderate symptom severity corresponds to a score of two or greater on the survey question (scores range from 0-4). Percentages were calculated by dividing the total number of participants experiencing a specific symptom by the total number of participants that filled out the survey for that time point.

## Discussion

The results from this hypothesis-generating pilot study demonstrate the novel nutraceutical, Qualia NAD+, boosts NAD+ levels an average of 74% in healthy, middle-aged males and females. Previous clinical trials reported modest increases (40 - 59%) in NAD+ using the dose of 300 mg/d nicotinamide riboside (NR) or nicotinamide mononucleotide (NMN), with larger increases requiring excess doses (1000 - 2000 mg/d)^41^. Our results suggest a blend of vitamins, nutrients and ingredients may be more effective in supporting overall NAD+ metabolism compared to NR or NMN alone given at similar doses. Improvements in quality of life measures were also noted only for males comparing baseline to day 28 in the overall and somatic categories. No other significant improvements in the quality of life measures were observed in the modified per-protocol (mPP) analysis, but the per-protocol (PP) analysis revealed those taking Qualia NAD+ exhibited significant improvements in energy/fatigue, emotional well-being, and social functioning at the end of the study.

While NAD⁺ blood results remained robust across both the mPP and intent-to-treat (ITT) analyses, the statistical significance observed in the survey data was no longer evident in the intent-to-treat analysis. This highlights the need for cautious interpretation, especially given there also were no corrections for multiple comparisons and the study experienced uneven participant attrition. It is also important to note this was a fully remote clinical trial, which may have contributed to the attrition. For this reason, the number of participants in the treatment group was small, limiting statistical power and generalizability of the findings.

Individual variability in responses was also noted, where NAD+ levels increased anywhere from 29% to 134% across all participants in the Qualia NAD+ group. Similarly, interindividual variability is reported across the literature and the impact of NAD+ boosting on tissue NAD+ levels is inconsistent^36,41^ Measurements of the NAD+ metabolome are also highly variable and complete characterization of the metabolome lacks consistency^50^. This variability in NAD+ metabolism across different populations has been attributed to multiple factors, including age, sex, and gut microbiota compositions^8,32,36,41,50^. This variance highlights the complexity of NAD+ regulation and the necessity for a therapeutic approach that considers multiple aspects of NAD+ metabolism in conjunction with improved modalities for characterizing the metabolome^6,51^. Moreover, changes in NAD+ metabolism enzymes are known to impact NAD+ levels and lifespan, offering a strong rationale for targeting them therapeutically.

Nicotinamide phosphoribosyltransferase (NAMPT), the rate-limiting step in the salvage pathway, reduces with aging in animals^2,52^ and humans^53-55^ across various tissues, but overexpression of the Nampt gene extends lifespan in mammalian cells^56,57^. Additionally, increased salvage pathway activity extends lifespan in yeast^58-60^. In fact, caloric restricted-mediated NAD+ boosting depends on the salvage pathway rather than the *de novo* pathway^59,61,62^ Nicotinate N-methyltransferases (NMNATs) responsible for synthesizing NAD+ have also been shown to decrease in some tissues with aging in animals^30^, while Nmnat3 overexpression in mice prevented age-related reductions in NAD levels^63^. Qualia NAD+ was formulated with additional nutrients and ingredients besides B3 vitamins to support NAMPT and NMNATs involved in the salvaging and synthesis of NAD+, respectively, as well as conversion to other molecules in the NAD metabolome (NADH, NADP+, and NADPH, respectively).

Emerging preclinical evidence demonstrates, for example, that resveratrol enhances NAMPT expression and activity, contributing to increased NAD+ levels ^64-66^. Caffeine has been shown to support the activity of nicotinamide mononucleotide adenylyltransferease (NMNAT) 2^67-69^, which is located predominantly in the brain and nervous system^70,71^, also leading to elevated NAD+ levels. Notably, co-administration of NR and caffeine results in greater increases in total NAD, NAD+, and NADH compared to NR or caffeine alone, suggesting their potentially additive or synergistic effects on NAD metabolism^68^. B vitamins support the kynurenine pathway and the energy production processes that are reliant upon the conversion of NAD+ to NADH^72-74^. Magnesium (with 71 other trace minerals) was also included to support adenosine triphosphate (ATP) synthesis, which is essential for the activity of most enzymes in the NAD+ biosynthesis pathway (**Figure 1**) as well as conversions among NAD+ and the other NAD metabolites. Since ATP must be complexed with magnesium to be biologically active, flux through the NAD+ metabolome is ultimately ATP and magnesium dependent^75-77^. In a premature aging mouse model, magnesium also promotes mitochondrial NADH oxidation (regeneration of NAD⁺) and enhances NADPH production, implicating a broader role of magnesium in modulating the NAD metabolome^78^. While these ingredients are mechanistically supported by preclinical evidence, further clinical research is needed to validate their effects on NAD+ metabolism and related health outcomes in humans.

Another primary limitation of our study is the fact that few other human clinical trials have measured NAD+ using the blood-spot assay^79^, though emerging research suggests that there is a good correlation between venous and fingerstick capillary blood NAD+ measurements^8^. This study also did not characterize the broader NAD+ metabolome or the NAD+/NADH redox ratio, which is increasingly more vital to understand the impacts of NAD+-boosting therapeutics^51^. While we focused on total NAD+ as an accessible endpoint, as it is the biomarker most measured in this field of research, we acknowledge that the NAD+/NADH ratio provides deeper insight into mitochondrial and cellular health. Lastly, due to the relatively short intervention period of 28 days and the absence of post-intervention follow-up, the durability of the observed effects remains unknown. While NAD+ boosting supplements such as NR and NMN have demonstrated a favorable safety profile in humans for up to 20 weeks of use ^36^, and nicotinic acid has been used safely for up to 6 years ^37^, long-term safety and use in specific vulnerable populations remain unclear, highlighting the need for more research. Future studies will be essential to evaluate the longer-term safety, outcomes or sustainability of beneficial effects over time. Given the current landscape of NAD+ research, it is important to continue to investigate relevant clinical and quality of life outcomes across a range of populations and with a variety of NAD+ metabolome biomarkers and measurement modalities.

The research for NAD+ boosting therapeutics in mitigating age-related disease and dysfunction is still in its early phases of exploration. Further, the impact of NAD+ boosting on improving healthy aging and healthspan is largely unknown. In this study, Qualia NAD+ increased whole blood NAD+ levels an average of 74% and this corresponded to some improvements in aging symptoms for males, but no improvements were observed for females. Given the study’s exploratory nature and limitations, including small sample size and potential underpowering, short duration, and limited generalizability, these results should be interpreted with caution. To advance this field, larger and longer-term randomized trials are needed to evaluate the potential of NAD+ boosting for improving healthspan in diverse populations.

## Data Availability

All data produced in the present study are available upon reasonable request to the authors.

## Acknowledgments

Not applicable.

## References

1. Harden A, Young WJ, Martin CJ. The alcoholic ferment of yeast-juice. Part II.—The coferment of yeast-juice. Proceedings of the Royal Society of London Series B, Containing Papers of a Biological Character. 1906;78(526):369–375. doi:doi:10.1098/rspb.1906.0070

2. Yoshino J, Mills KF, Yoon MJ, Imai S. Nicotinamide mononucleotide, a key NAD(+) intermediate, treats the pathophysiology of diet- and age-induced diabetes in mice. Cell Metab. Oct 5 2011;14(4):528–36. doi:10.1016/j.cmet.2011.08.014

3. Camacho-Pereira J, Tarragó MG, Chini CCS, et al. CD38 Dictates Age-Related NAD Decline and Mitochondrial Dysfunction through an SIRT3-Dependent Mechanism. Cell Metab. Jun 14 2016;23(6):1127–1139. doi:10.1016/j.cmet.2016.05.006

4. Massudi H, Grant R, Braidy N, Guest J, Farnsworth B, Guillemin GJ. Age-Associated Changes In Oxidative Stress and NAD+ Metabolism In Human Tissue. PLOS ONE. 2012;7(7):e42357. doi:10.1371/journal.pone.0042357

5. Zhu X-H, Lu M, Lee B-Y, Ugurbil K, Chen W. In vivo NAD assay reveals the intracellular NAD contents and redox state in healthy human brain and their age dependences. Proceedings of the National Academy of Sciences. 2015/03/03 2015;112(9):2876-2881. doi:10.1073/pnas.1417921112

6. Clement J, Wong M, Poljak A, Sachdev P, Braidy N. The Plasma NAD+ Metabolome Is Dysregulated in “Normal” Aging. Rejuvenation Research. 2019/04/01 2018;22(2):121-130. doi:10.1089/rej.2018.2077

7. Pospieszna B, Kusy K, Slominska EM, Zieliński J, Ciekot-Sołtysiak M. Erythrocyte nicotinamide adenine dinucleotide concentration is enhanced by systematic sports participation. BMC Sports Sci Med Rehabil. Oct 15 2024;16(1):216. doi:10.1186/s13102-024-00999-y

8. Wang P, Chen M, Hou Y, et al. Fingerstick blood assay maps real-world NAD(+) disparity across gender and age. Aging Cell. Oct 2023;22(10):e13965. doi:10.1111/acel.13965

9. Janssens GE, Grevendonk L, Perez RZ, et al. Healthy aging and muscle function are positively associated with NAD(+) abundance in humans. Nat Aging. Mar 2022;2(3):254–263. doi:10.1038/s43587-022-00174-3

10. Guo R, Yang S, Wiesner HM, et al. Mapping intracellular NAD content in entire human brain using phosphorus-31 MR spectroscopic imaging at 7 Tesla. Original Research. Frontiers in Neuroscience. 2024-June-07 2024;18doi:10.3389/fnins.2024.1389111

11. Kataura T, Sedlackova L, Otten EG, et al. Autophagy promotes cell survival by maintaining NAD levels. Developmental Cell. 2022;57(22):2584–2598.e11. doi:10.1016/j.devcel.2022.10.008

12. Mills KF, Yoshida S, Stein LR, et al. Long-Term Administration of Nicotinamide Mononucleotide Mitigates Age-Associated Physiological Decline in Mice. Cell Metab. Dec 13 2016;24(6):795–806. doi:10.1016/j.cmet.2016.09.013

13. Diguet N, Trammell SAJ, Tannous C, et al. Nicotinamide Riboside Preserves Cardiac Function in a Mouse Model of Dilated Cardiomyopathy. Circulation. 2018/05/22 2018;137(21):2256-2273. doi:10.1161/CIRCULATIONAHA.116.026099

14. Hou Y, Dan X, Babbar M, et al. Ageing as a risk factor for neurodegenerative disease. Nature Reviews Neurology. 2019/10/01 2019;15(10):565-581. doi:10.1038/s41582-019-0244-7

15. Yulug B, Altay O, Li X, et al. Combined Metabolic Activators Improve Cognitive Functions without Altering Motor Scores in Parkinson’s Disease. medRxiv. 2021:2021.07.28.21261293. doi:10.1101/2021.07.28.21261293

16. Yulug B, Altay O, Li X, et al. Combined Metabolic Activators Improves Cognitive Functions in Alzheimer’s Disease. medRxiv. 2021:2021.07.14.21260511. doi:10.1101/2021.07.14.21260511

17. Verdin E. NAD⁺ in aging, metabolism, and neurodegeneration. Science. Dec 4 2015;350(6265):1208–13. doi:10.1126/science.aac4854

18. Cantó C, Sauve AA, Bai P. Crosstalk between poly(ADP-ribose) polymerase and sirtuin enzymes. Mol Aspects Med. Dec 2013;34(6):1168–201. doi:10.1016/j.mam.2013.01.004

19. Xie N, Zhang L, Gao W, et al. NAD(+) metabolism: pathophysiologic mechanisms and therapeutic potential. Signal Transduct Target Ther. Oct 7 2020;5(1):227. doi:10.1038/s41392-020-00311-7

20. Piedra-Quintero ZL, Wilson Z, Nava P, Guerau-de-Arellano M. CD38: An Immunomodulatory Molecule in Inflammation and Autoimmunity. Review. Frontiers in Immunology. 2020-November-30 2020;Volume 11 - 2020doi:10.3389/fimmu.2020.597959

21. Lautrup S, Sinclair DA, Mattson MP, Fang EF. NAD(+) in Brain Aging and Neurodegenerative Disorders. Cell Metab. Oct 1 2019;30(4):630–655. doi:10.1016/j.cmet.2019.09.001

22. Bai P, Cantó C, Oudart H, et al. PARP-1 inhibition increases mitochondrial metabolism through SIRT1 activation. Cell Metab. Apr 6 2011;13(4):461–468. doi:10.1016/j.cmet.2011.03.004

23. Mottahedeh J, Haffner MC, Grogan TR, et al. CD38 is methylated in prostate cancer and regulates extracellular NAD(). Cancer Metab. 2018;6:13. doi:10.1186/s40170-018-0186-3

24. Imai S. The NAD World: a new systemic regulatory network for metabolism and aging--Sirt1, systemic NAD biosynthesis, and their importance. Cell Biochem Biophys. 2009;53(2):65–74. doi:10.1007/s12013-008-9041-4

25. Chini EN. CD38 as a regulator of cellular NAD: a novel potential pharmacological target for metabolic conditions. Curr Pharm Des. 2009;15(1):57–63. doi:10.2174/138161209787185788

26. Hurtado-Bagès S, Knobloch G, Ladurner AG, Buschbeck M. The taming of PARP1 and its impact on NAD(+) metabolism. Mol Metab. Aug 2020;38:100950. doi:10.1016/j.molmet.2020.01.014

27. Mouchiroud L, Houtkooper RH, Moullan N, et al. The NAD(+)/Sirtuin Pathway Modulates Longevity through Activation of Mitochondrial UPR and FOXO Signaling. Cell. Jul 18 2013;154(2):430–41. doi:10.1016/j.cell.2013.06.016

28. Chini EN, Chini CCS, Espindola Netto JM, de Oliveira GC, van Schooten W. The Pharmacology of CD38/NADase: An Emerging Target in Cancer and Diseases of Aging. Trends Pharmacol Sci. Apr 2018;39(4):424–436. doi:10.1016/j.tips.2018.02.001

29. Ubaida-Mohien C, Lyashkov A, Gonzalez-Freire M, et al. Discovery proteomics in aging human skeletal muscle finds change in spliceosome, immunity, proteostasis and mitochondria. Elife. Oct 23 2019;8doi:10.7554/eLife.49874

30. McReynolds MR, Chellappa K, Baur JA. Age-related NAD(+) decline. Exp Gerontol. Feb 22 2020;134:110888. doi:10.1016/j.exger.2020.110888

31. Fang EF, Lautrup S, Hou Y, et al. NAD(+) in Aging: Molecular Mechanisms and Translational Implications. Trends Mol Med. Oct 2017;23(10):899–916. doi:10.1016/j.molmed.2017.08.001

32. Iqbal T, Nakagawa T. The therapeutic perspective of NAD(+) precursors in age-related diseases. Biochem Biophys Res Commun. Apr 2 2024;702:149590. doi:10.1016/j.bbrc.2024.149590

33. Poljšak B, Kovač V, Špalj S, Milisav I. The Central Role of the NAD+ Molecule in the Development of Aging and the Prevention of Chronic Age-Related Diseases: Strategies for NAD+ Modulation. Int J Mol Sci. Feb 3 2023;24(3)doi:10.3390/ijms24032959

34. Gilmour BC, Gudmundsrud R, Frank J, et al. Targeting NAD(+) in translational research to relieve diseases and conditions of metabolic stress and ageing. Mech Ageing Dev. Mar 2020;186:111208. doi:10.1016/j.mad.2020.111208

35. Conze D, Brenner C, Kruger CL. Safety and Metabolism of Long-term Administration of NIAGEN (Nicotinamide Riboside Chloride) in a Randomized, Double-Blind, Placebo-controlled Clinical Trial of Healthy Overweight Adults. Scientific Reports. 2019/07/05 2019;9(1):9772. doi:10.1038/s41598-019-46120-z

36. Freeberg KA, Udovich CC, Martens CR, Seals DR, Craighead DH. Dietary Supplementation With NAD+-Boosting Compounds in Humans: Current Knowledge and Future Directions. J Gerontol A Biol Sci Med Sci. Dec 1 2023;78(12):2435–2448. doi:10.1093/gerona/glad106

37. Zeman M, Vecka M, Perlík F, et al. Niacin in the Treatment of Hyperlipidemias in Light of New Clinical Trials: Has Niacin Lost its Place? Med Sci Monit. Jul 25 2015;21:2156–62. doi:10.12659/msm.893619

38. Ying W. NAD+/NADH and NADP+/NADPH in Cellular Functions and Cell Death: Regulation and Biological Consequences. Antioxidants & Redox Signaling. 2008/02/01 2007;10(2):179-206. doi:10.1089/ars.2007.1672

39. Schwarzmann L, Pliquett RU, Simm A, Bartling B. Sex-related differences in human plasma NAD+/NADH levels depend on age. Biosci Rep. Jan 29 2021;41(1)doi:10.1042/bsr20200340

40. Xue Y, Shamp T, Nagana Gowda GA, Crabtree M, Bagchi D, Raftery D. A Combination of Nicotinamide and D-Ribose (RiaGev) Is Safe and Effective to Increase NAD(+) Metabolome in Healthy Middle-Aged Adults: A Randomized, Triple-Blind, Placebo-Controlled, Cross-Over Pilot Clinical Trial. Nutrients. May 26 2022;14(11)doi:10.3390/nu14112219

41. Henderson JD, Quigley SNZ, Chachra SS, Conlon N, Ford D. The use of a systems approach to increase NAD+ in human participants. npj Aging. 2024/02/01 2024;10(1):7. doi:10.1038/s41514-023-00134-0

42. Braidy N, Villalva MD, Grant R. NADomics: Measuring NAD(+) and Related Metabolites Using Liquid Chromatography Mass Spectrometry. Life (Basel). May 31 2021;11(6)doi:10.3390/life11060512

43. Smith S, Ellgass M, Park A, et al. P053: Understanding the measurement of nicotinamide adenine dinucleotide (NAD+) from dried blood spots through a population study. Genetics in Medicine Open. 2024;2doi:10.1016/j.gimo.2024.100930

44. Matsuyama R, Omata T, Kageyama M, Nakajima R, Kanou M, Yamana K. Stabilization and quantitative measurement of nicotinamide adenine dinucleotide in human whole blood using dried blood spot sampling. Anal Bioanal Chem. Feb 2023;415(5):775–785. doi:10.1007/s00216-022-04469-7

45. Airhart SE, Shireman LM, Risler LJ, et al. An open-label, non-randomized study of the pharmacokinetics of the nutritional supplement nicotinamide riboside (NR) and its effects on blood NAD+ levels in healthy volunteers. PLoS One. 2017;12(12):e0186459. doi:10.1371/journal.pone.0186459

46. Smith S, Fox E, Euro L, et al. eP032: Measurement of Nicotinamide Adenine Dinucleotide (NAD+) from dried blood spot cards. Genetics in Medicine. 2022;24(3):S21–S22. doi:10.1016/j.gim.2022.01.070

47. Heinemann LA. Aging Males’ Symptoms scale: a standardized instrument for the practice. J Endocrinol Invest. 2005;28(11 Suppl Proceedings):34-8.

48. Hays RD, Sherbourne CD, Mazel RM. The RAND 36-Item Health Survey 1.0. Health Econ. Oct 1993;2(3):217–27. doi:10.1002/hec.4730020305

49. Ware JE, Jr., Sherbourne CD. The MOS 36-item short-form health survey (SF-36). I. Conceptual framework and item selection. Med Care. Jun 1992;30(6):473–83.

50. Gindri IM, Ferrari G, Pinto LPS, et al. Evaluation of safety and effectiveness of NAD in different clinical conditions: a systematic review. Am J Physiol Endocrinol Metab. Apr 1 2024;326(4):E417–e427. doi:10.1152/ajpendo.00242.2023

51. Migaud ME, Ziegler M, Baur JA. Regulation of and challenges in targeting NAD(+) metabolism. Nat Rev Mol Cell Biol. Oct 2024;25(10):822–840. doi:10.1038/s41580-024-00752-w

52. Jadeja RN, Powell FL, Jones MA, et al. Loss of NAMPT in aging retinal pigment epithelium reduces NAD(+) availability and promotes cellular senescence. Aging (Albany NY). Jun 12 2018;10(6):1306–1323. doi:10.18632/aging.101469

53. de Guia RM, Agerholm M, Nielsen TS, et al. Aerobic and resistance exercise training reverses age-dependent decline in NAD(+) salvage capacity in human skeletal muscle. Physiol Rep. Jul 2019;7(12):e14139. doi:10.14814/phy2.14139

54. Yoshida M, Satoh A, Lin JB, et al. Extracellular Vesicle-Contained eNAMPT Delays Aging and Extends Lifespan in Mice. Cell Metab. Aug 6 2019;30(2):329–342.e5. doi:10.1016/j.cmet.2019.05.015

55. Zhou CC, Yang X, Hua X, et al. Hepatic NAD(+) deficiency as a therapeutic target for non-alcoholic fatty liver disease in ageing. Br J Pharmacol. Aug 2016;173(15):2352–68. doi:10.1111/bph.13513

56. Borradaile NM, Pickering JG. Nicotinamide phosphoribosyltransferase imparts human endothelial cells with extended replicative lifespan and enhanced angiogenic capacity in a high glucose environment. Aging Cell. Apr 2009;8(2):100–12. doi:10.1111/j.1474-9726.2009.00453.x

57. van der Veer E, Ho C, O’Neil C, et al. Extension of human cell lifespan by nicotinamide phosphoribosyltransferase. J Biol Chem. Apr 13 2007;282(15):10841–5. doi:10.1074/jbc.C700018200

58. Anderson RM, Bitterman KJ, Wood JG, Medvedik O, Sinclair DA. Nicotinamide and PNC1 govern lifespan extension by calorie restriction in Saccharomyces cerevisiae. Nature. 2003/05/01 2003;423(6936):181-185. doi:10.1038/nature01578

59. Lin SJ, Defossez PA, Guarente L. Requirement of NAD and SIR2 for life-span extension by calorie restriction in Saccharomyces cerevisiae. Science. Sep 22 2000;289(5487):2126–8. doi:10.1126/science.289.5487.2126

60. Anderson RM, Bitterman KJ, Wood JG, et al. Manipulation of a Nuclear NAD+ Salvage Pathway Delays Aging without Altering Steady-state NAD+ Levels*. Journal of Biological Chemistry. 2002/05/24/ 2002;277(21):18881-18890. 10.1074/jbc.M111773200

61. Lin SJ, Ford E, Haigis M, Liszt G, Guarente L. Calorie restriction extends yeast life span by lowering the level of NADH. Genes Dev. Jan 1 2004;18(1):12–6. doi:10.1101/gad.1164804

62. Lin SJ, Kaeberlein M, Andalis AA, et al. Calorie restriction extends Saccharomyces cerevisiae lifespan by increasing respiration. Nature. Jul 18 2002;418(6895):344–8. doi:10.1038/nature00829

63. Gulshan M, Yaku K, Okabe K, et al. Overexpression of Nmnat3 efficiently increases NAD and NGD levels and ameliorates age-associated insulin resistance. Aging Cell. Aug 2018;17(4):e12798. doi:10.1111/acel.12798

64. Schuster S, Penke M, Gorski T, et al. Resveratrol Differentially Regulates NAMPT and SIRT1 in Hepatocarcinoma Cells and Primary Human Hepatocytes. PLOS ONE. 2014;9(3):e91045. doi:10.1371/journal.pone.0091045

65. Lan F, Weikel KA, Cacicedo JM, Ido Y. Resveratrol-Induced AMP-Activated Protein Kinase Activation Is Cell-Type Dependent: Lessons from Basic Research for Clinical Application. Nutrients. Jul 14 2017;9(7)doi:10.3390/nu9070751

66. Huang P, Riordan SM, Heruth DP, Grigoryev DN, Zhang LQ, Ye SQ. A critical role of nicotinamide phosphoribosyltransferase in human telomerase reverse transcriptase induction by resveratrol in aortic smooth muscle cells. Oncotarget. May 10 2015;6(13):10812–24. doi:10.18632/oncotarget.3580

67. Zwilling M, Theiss C, Matschke V. Caffeine and NAD(+) Improve Motor Neural Integrity of Dissociated Wobbler Cells In Vitro. Antioxidants (Basel). May 27 2020;9(6)doi:10.3390/antiox9060460

68. Ryu WI, Shen M, Lee Y, et al. Nicotinamide riboside and caffeine partially restore diminished NAD availability but not altered energy metabolism in Alzheimer’s disease. Aging Cell. Jul 2022;21(7):e13658. doi:10.1111/acel.13658

69. Ali YO, Bradley G, Lu H-C. Screening with an NMNAT2-MSD platform identifies small molecules that modulate NMNAT2 levels in cortical neurons. Scientific Reports. 2017/03/07 2017;7(1):43846. doi:10.1038/srep43846

70. Mayer PR, Huang N, Dewey CM, Dries DR, Zhang H, Yu G. Expression, localization, and biochemical characterization of nicotinamide mononucleotide adenylyltransferase 2. J Biol Chem. Dec 17 2010;285(51):40387–96. doi:10.1074/jbc.M110.178913

71. Yaku K, Okabe K, Nakagawa T. NAD metabolism: Implications in aging and longevity. Ageing Res Rev. Nov 2018;47:1–17. doi:10.1016/j.arr.2018.05.006

72. Hanna M, Jaqua E, Nguyen V, Clay J. B Vitamins: Functions and Uses in Medicine. Perm J. Jun 29 2022;26(2):89-97. doi:10.7812/tpp/21.204

73. Hrubša M, Siatka T, Nejmanová I, et al. Biological Properties of Vitamins of the B-Complex, Part 1: Vitamins B(1), B(2), B(3), and B(5). Nutrients. Jan 22 2022;14(3)doi:10.3390/nu14030484

74. Majewski M, Kozlowska A, Thoene M, Lepiarczyk E, Grzegorzewski WJ. Overview of the role of vitamins and minerals on the kynurenine pathway in health and disease. J Physiol Pharmacol. Feb 2016;67(1):3–19.

75. Lau C, Niere M, Ziegler M. The NMN/NaMN adenylyltransferase (NMNAT) protein family. Front Biosci (Landmark Ed). Jan 1 2009;14(2):410–31. doi:10.2741/3252

76. She J, Sheng R, Qin ZH. Pharmacology and Potential Implications of Nicotinamide Adenine Dinucleotide Precursors. Aging Dis. Dec 2021;12(8):1879–1897. doi:10.14336/ad.2021.0523

77. Storer AC, Cornish-Bowden A. Concentration of MgATP2- and other ions in solution. Calculation of the true concentrations of species present in mixtures of associating ions. Biochem J. Oct 1 1976;159(1):1–5. doi:10.1042/bj1590001

78. Villa-Bellosta R. Dietary magnesium supplementation improves lifespan in a mouse model of progeria. EMBO Mol Med. Oct 7 2020;12(10):e12423. doi:10.15252/emmm.202012423

79. Hawkins J, Idoine R, Kwon J, et al. Randomized, placebo-controlled, pilot clinical study evaluating acute Niagen®+ IV and NAD+ IV in healthy adults. medRxiv. 2024:2024.06.06.24308565. doi:10.1101/2024.06.06.24308565

